# Breastfeeding Practices Among Mothers During COVID-19 in India

**DOI:** 10.1101/2021.08.06.21261582

**Authors:** Ajoke Akinola Akinola, N. H. Simon, Nasteha Abdikadir Mohamed

## Abstract

**Introduction:** The Covid-19 pandemic is disrupting normal life globally. The COVID-19 pandemic is an emerging concern regarding the potential effects during breastfeeding. The aim of this study was to conduct a systemic review of mother-to -child transmission of COVID-19 during breastfeeding.

**Method:** This study systematically searched electronic databases; google scholar, PubMed, Medline, up December 2020. The study was included studies relevant to transmission breast milk and respiratory droplets during breastfeeding of mothers with COVID-19 positive. To identify the quality of data, prism standard was used and Strobe checklist scale.

**Result:** A total of 3160 records were identified in this systemic review with eight relevant studies involving 159 mothers (63 mothers with COVID-19 positive, 55 of their breast milk samples tested negative for the-Covid-19. Twenty-one breast milk samples from 8 women tested positive for Covid-19. Of 73 infants were born to mothers with COVID-19 at the time of delivery. Two infants tested positive for Covid-19. The average mother-child separation time was 36•7 ± 21•1 days among mothers confirmed with COVID-19. Out of 22 mothers, ((37.5%) chose to breastfeed their babies after confirm covid-19 positive.

**Conclusion:** This study shown that breastfeeding practices were extremely impacted during the COVID-19 epidemic among both confirmed positive cases and suspected mothers. However, the risk of mother-to-infant transmission of Covid-19 vertically or horizontally, in the perinatal period is very low.

## Background

The coronavirus COVID-19 outbreak, caused by the extreme respiratory syndrome coronavirus 2 (SARS-CoV-2), was originated from the first December 2019 from Wuhan city in China (Poreddi, 2020). The first case of covid-2019 was identified in India on 30 January 2020, in a student who has returned home from Wuhan University in China (1, 2021). Since its outbreak covid-2019 has disturbing all aspect of life in around the world. The transmission of virus happens through respiratory droplets most notably, in close contact, crowed and touching and due to this, global level health measures have proposed to contain its spread that were the implementation of lockdowns, confinement, and social distancing (2, 2020). These global health measures to control the spread of covid-2019 have led the separation of mothers and their infants after birth, especially, suspected positive cases of covid-2019 or even confirmed positive mothers that resulted also separating mother-baby contact included breastfeeding. The covid-2019 global health measures including separation of the mother-infant has great negative effect on breastfeeding and child cares.

Breastfeeding practice has been healthier source for infants and it is the most well-known recommended method for infant feeding. Breastfeeding has two types, (a) Optimal breastfeeding and (b) exclusive breastfeeding. Optimal breastfeeding is the first crucial care for the infant within one hour after birth including; young feeding practices from birth to 2 years to fulfil the rights of infants to attain highest attainable standards of health (3, 2020). Exclusive breastfeeding on other hand, is the first six months, the adequate and suitable feeding at completion of six months (4, 2019). According to the guidelines by World Health Organisation and Government of India, optimal breastfeeding should be initiated within one hour of birth, and also exclusive breastfeeding for the first six months. WHO, further suggested that Mothers who tested positive, there are three possible considerations about care of infant that included; the use of donor milk, the use of expressed breast milk, and breastfeeding with precautions. advised all mothers to take necessary measures for the well-being and health of infant that including wearing face mask, washing hands frequently before making any contact to the baby (5, 2021).

Breastfeeding practice among mothers during covid 19

Breastfeeding has a great role to play in preventing an extreme infant life infectious disease during covid-2019. India is the highest country of the new-born mortality rate and Covid-19 worsened the situation that has posed many challenges to the provision of new-born nutrition (breastfeeding and maternal support). Nearly 25 million births, (43% (11 million) are not able to get breastfed within the first hour (6, 2019). Lack of inadequate breastfeeding has resulted the death of 100,00 with preventable disease diarrhoea and pneumonia. The covid19 is also worsened the lack of child care issues in India because separating covid19 positive mothers their babies have posed a new challenge to breastfeeding and new-born mortality rate. These new covid19 challenge caused that mother are worrying about the safety of feeding their infant due to health measures to separate mothers and infant in which has great impact of Indian mother’s mental health and safety of the infant.

The coronavirus covid19 respiratory syndrome virus 2 (SARS-CoV-2), has posed a huge challenge for providing necessary care to mothers, breastfeeding, and infants (7, 2020). Because breastfeeding has become susceptible concern about practicing skin-to skin contact that resulted separation of mothers, babies and inadequate care of infants. This is because has raised many fears on possible vertical transmission through breastfeeding and is presently main concern of new mothers and the evidence of vertical transmission is still conflicting (8, 2021). Therefore, this study aims to examine breastfeeding practice among mothers during Covid19 in India.

## METHODOLOGY

### Search strategy

To gather available literature for this systematic review a search was conducted through PubMed, Google Scholar and Medline databases, up December 2020. To identify the relevant articles, the search reviewed all related articles of Coronavirus, COVID-19, 2019-ncov, SARS-cov-2, of Breastfeeding Practice Among Mothers During Covid-19.

**Table 1:**
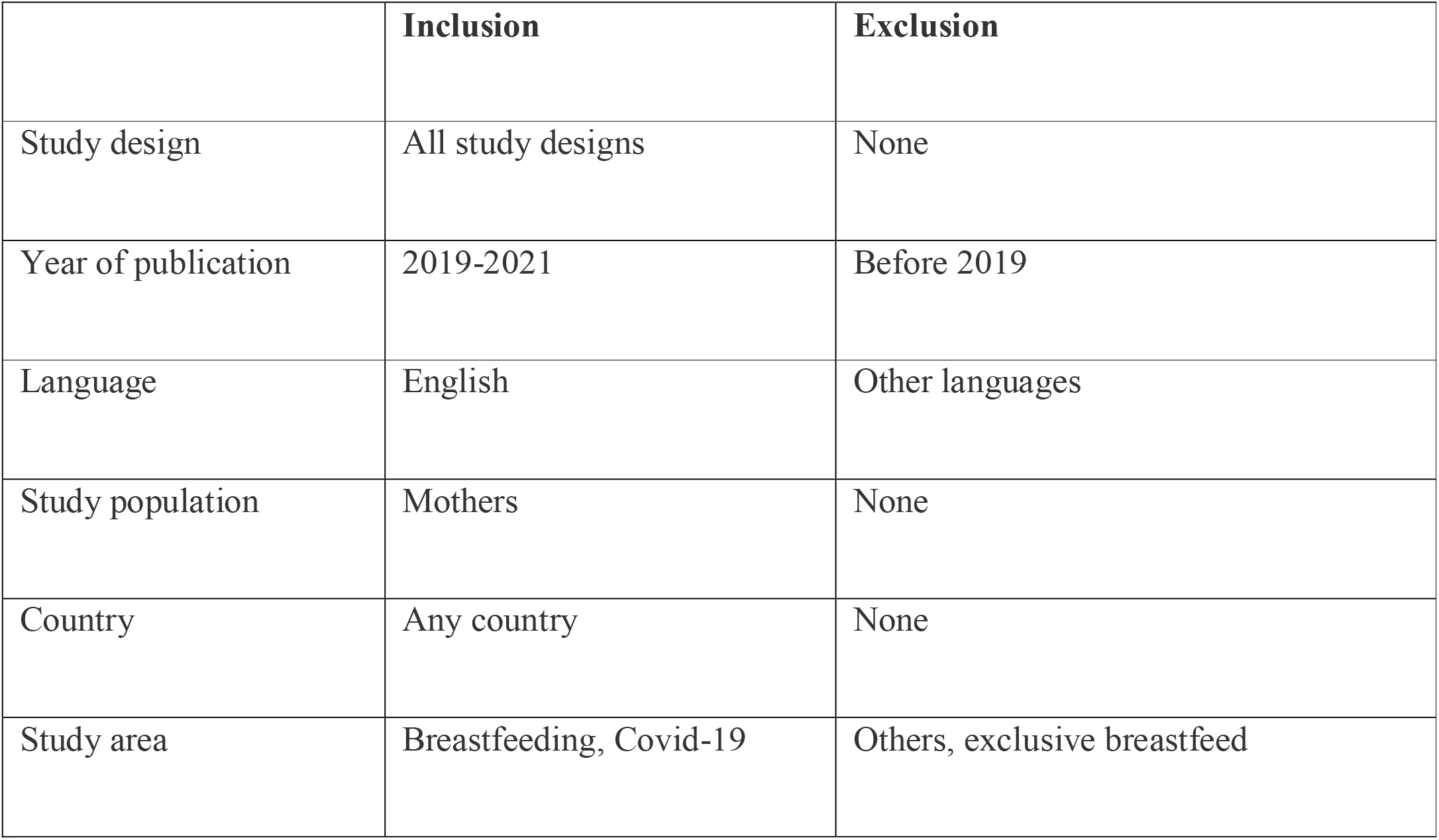
Inclusion and exclusion criteria.

**Table 2:**
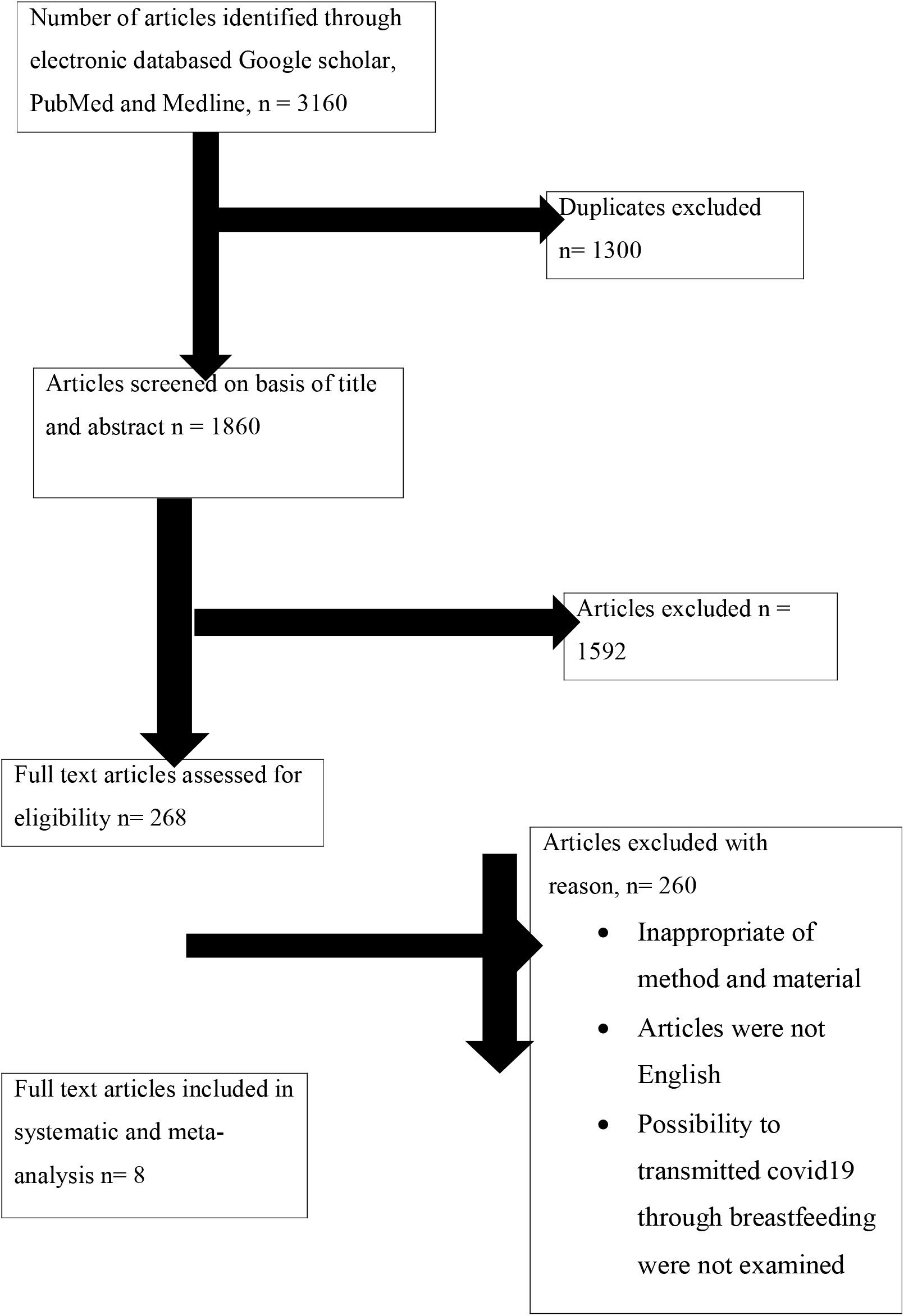
Prisma Table.

### Literature Review

Various studies and empirical literature pertinent to the study will revie

**Table 3:**
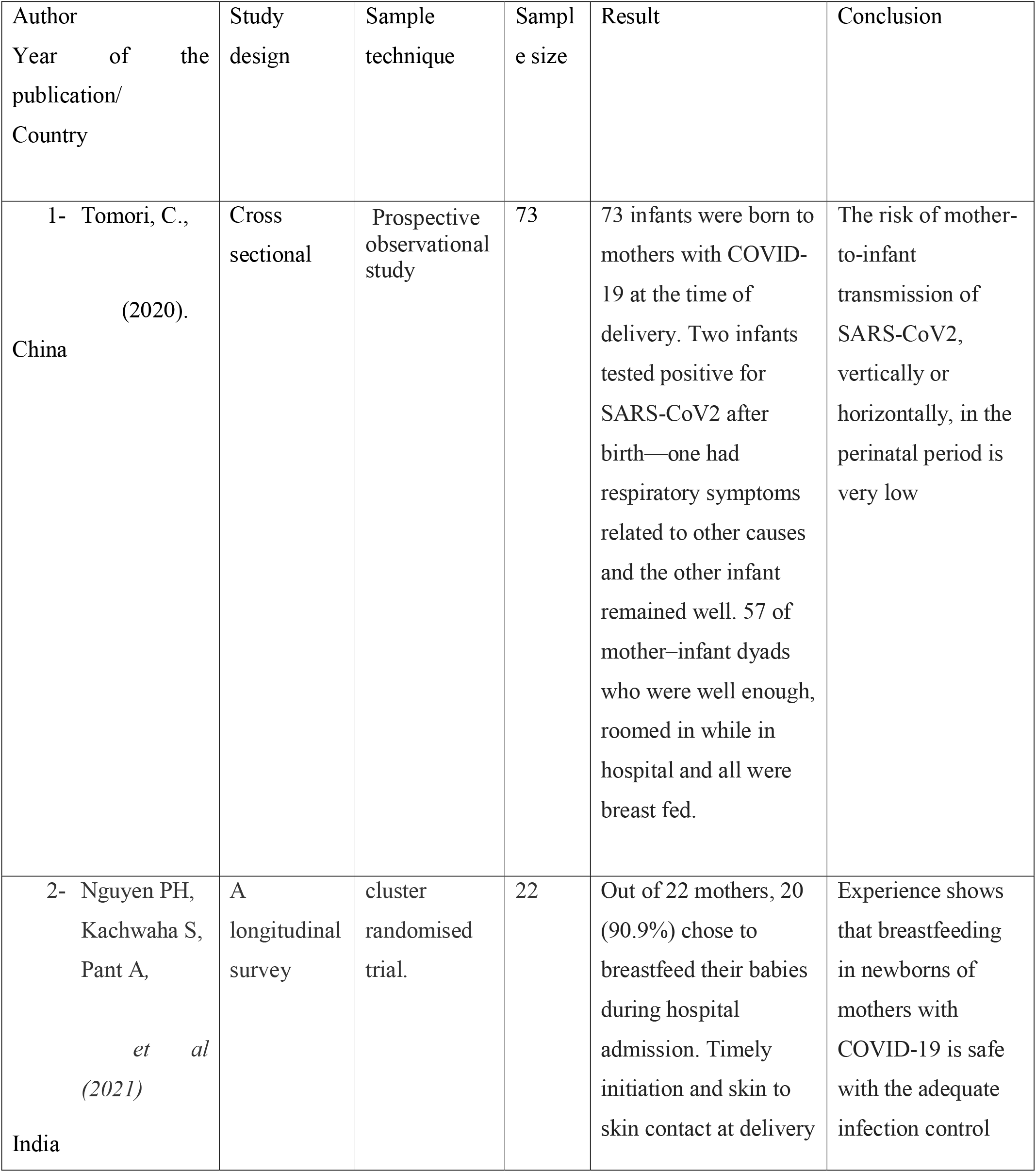
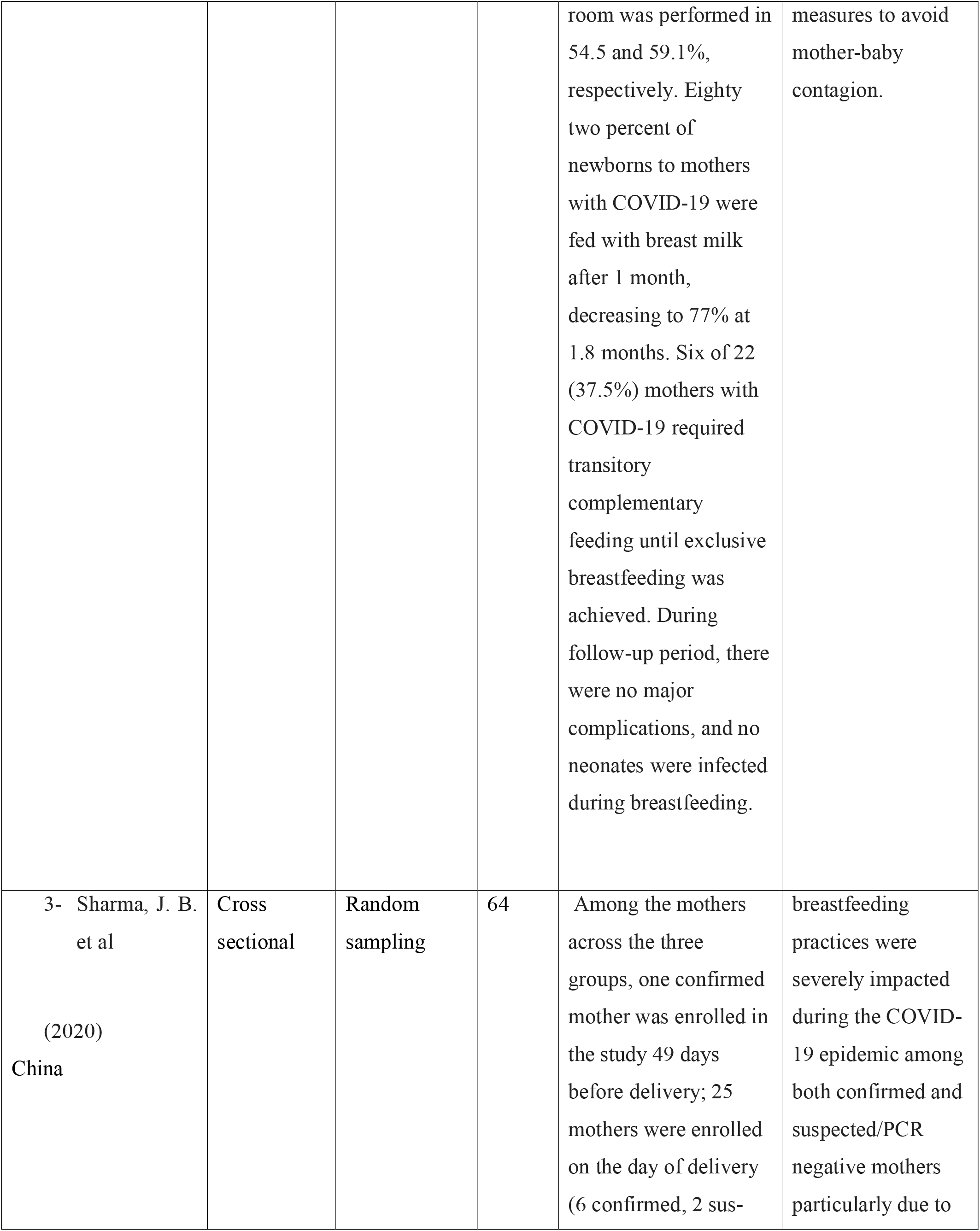
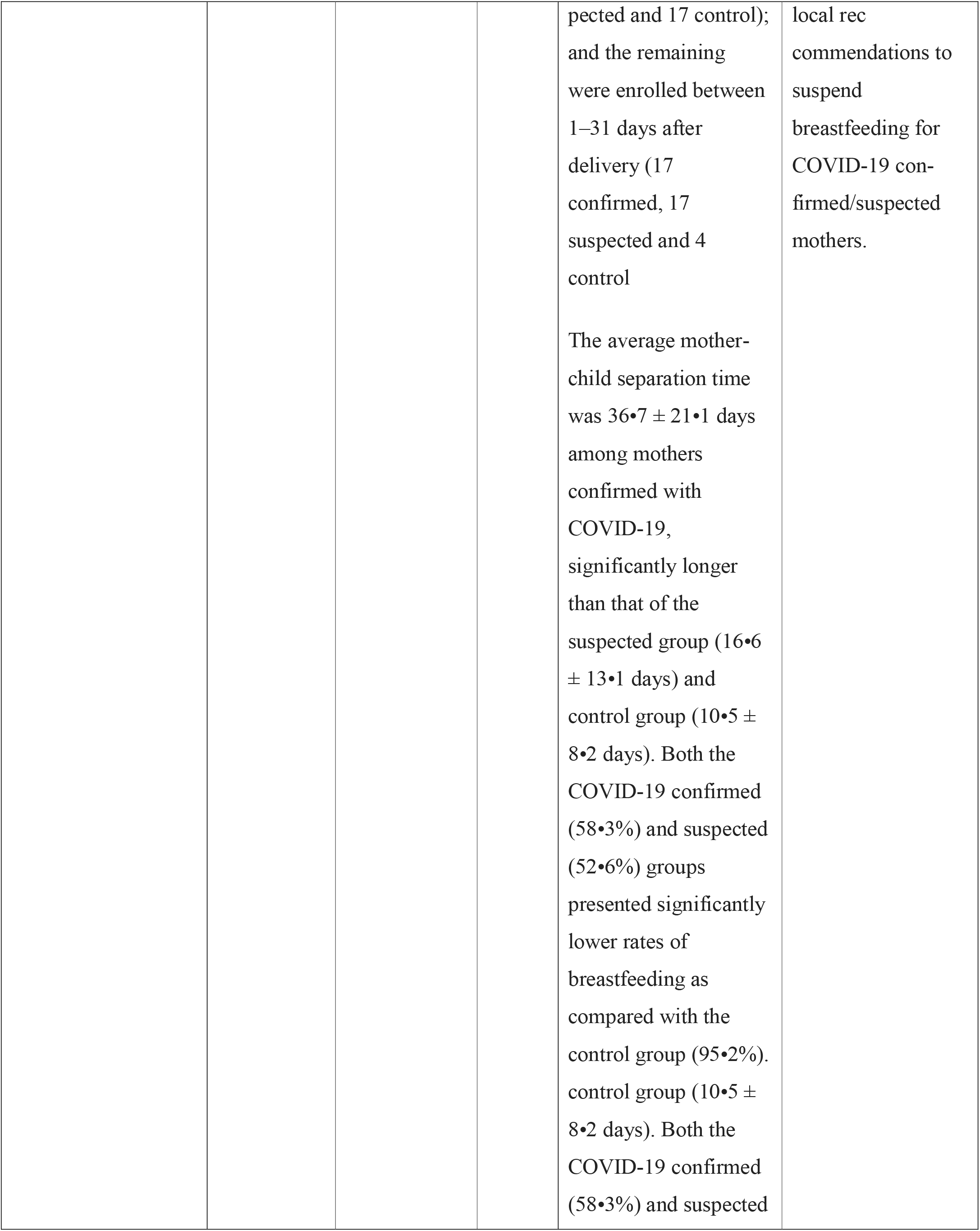
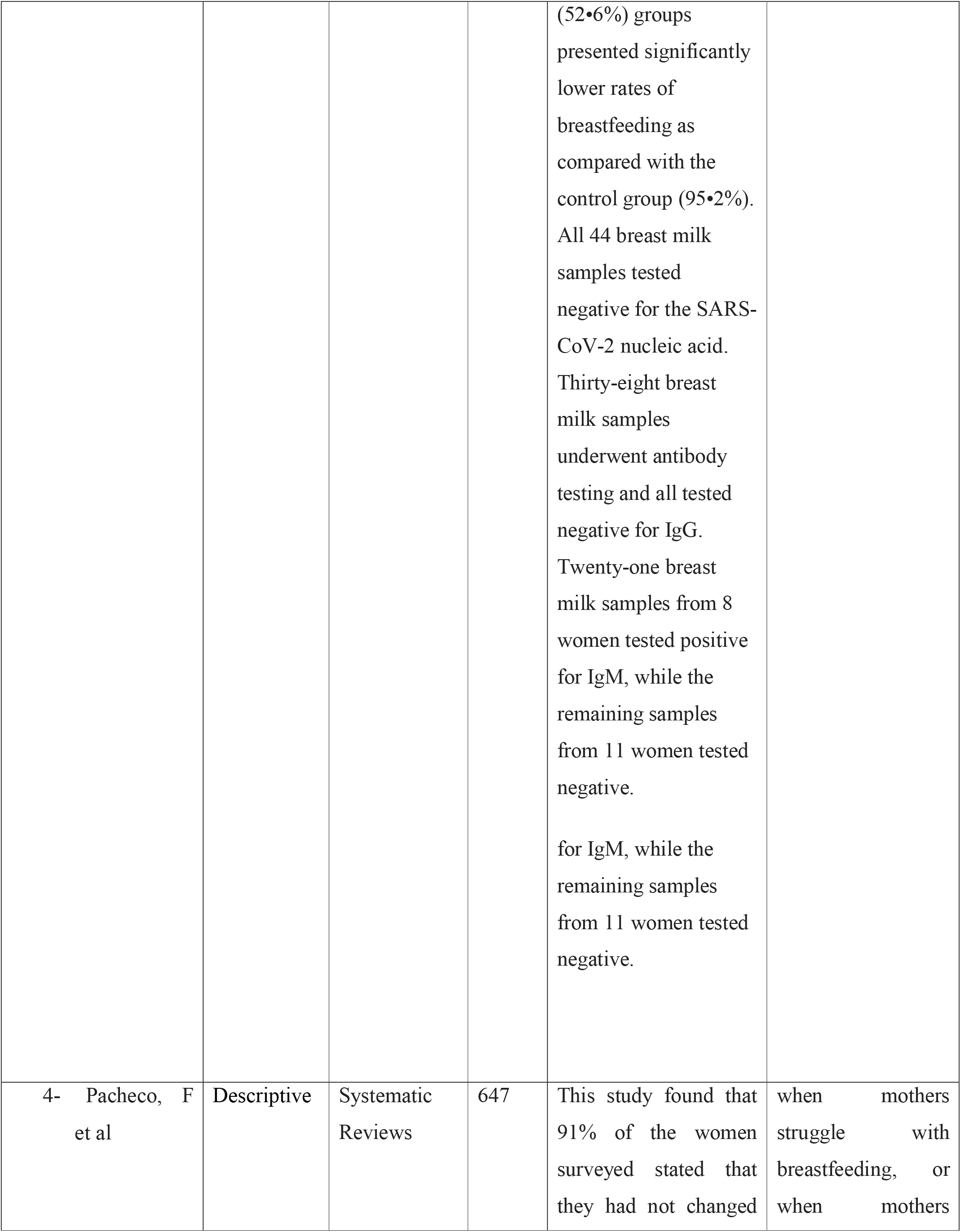
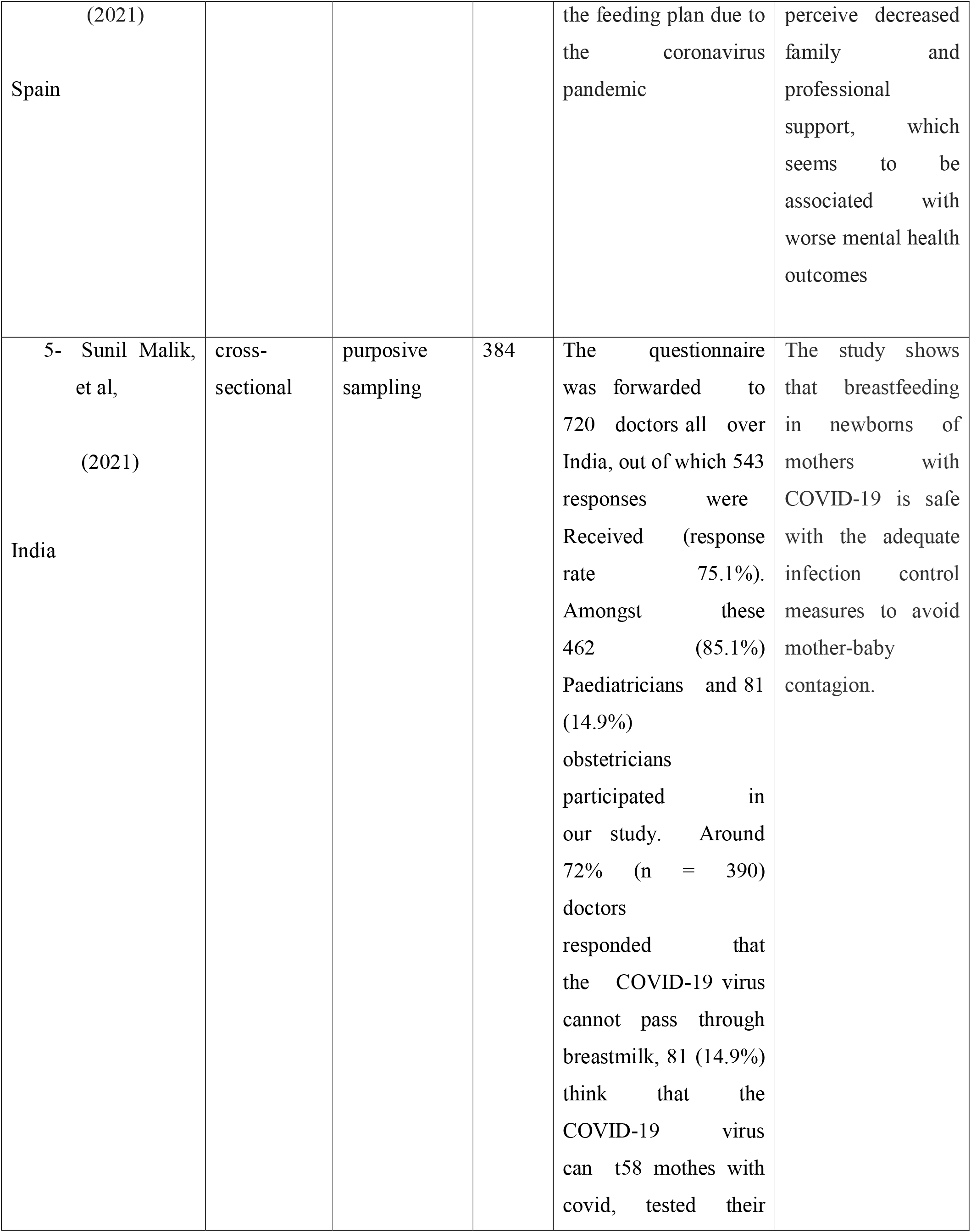
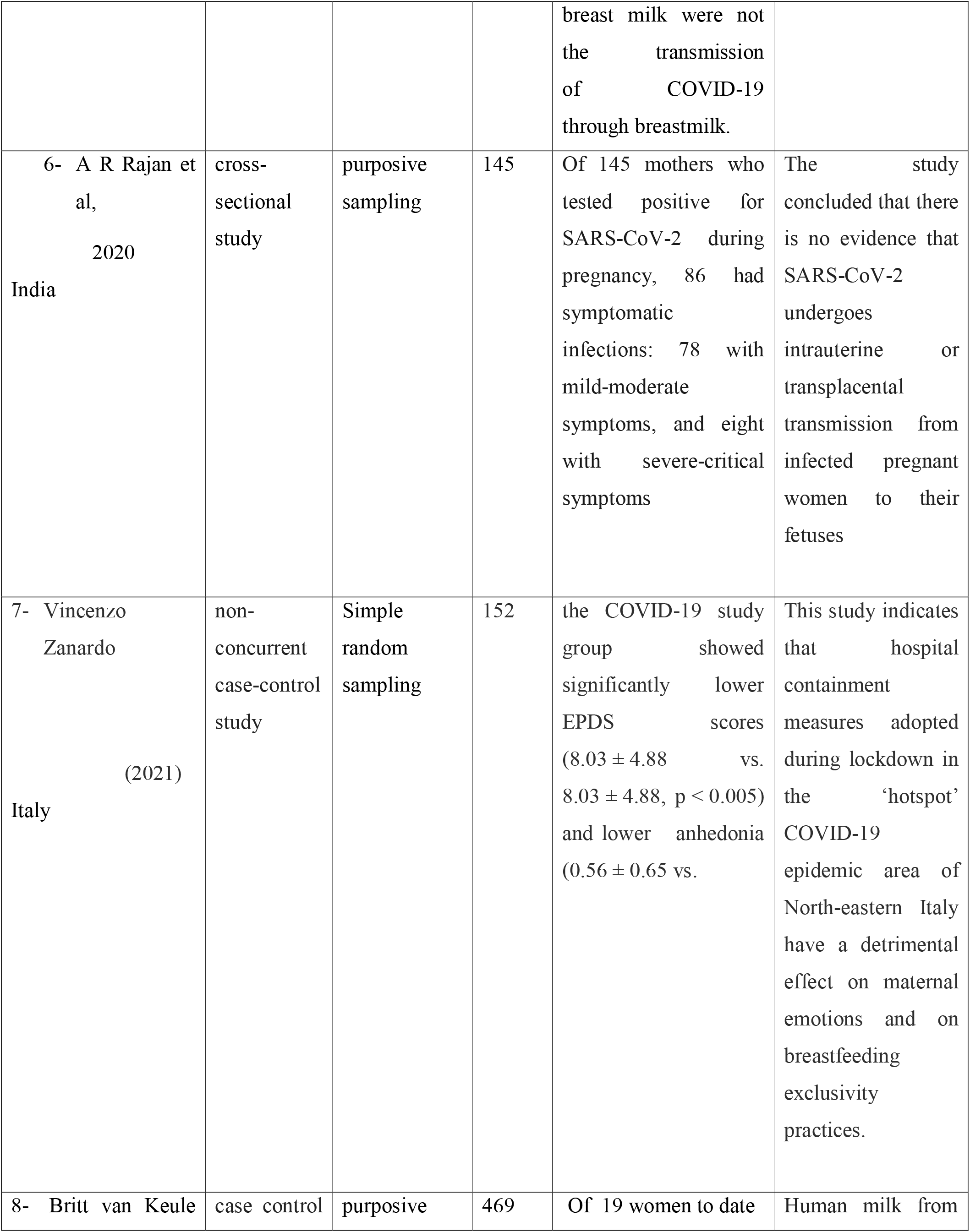
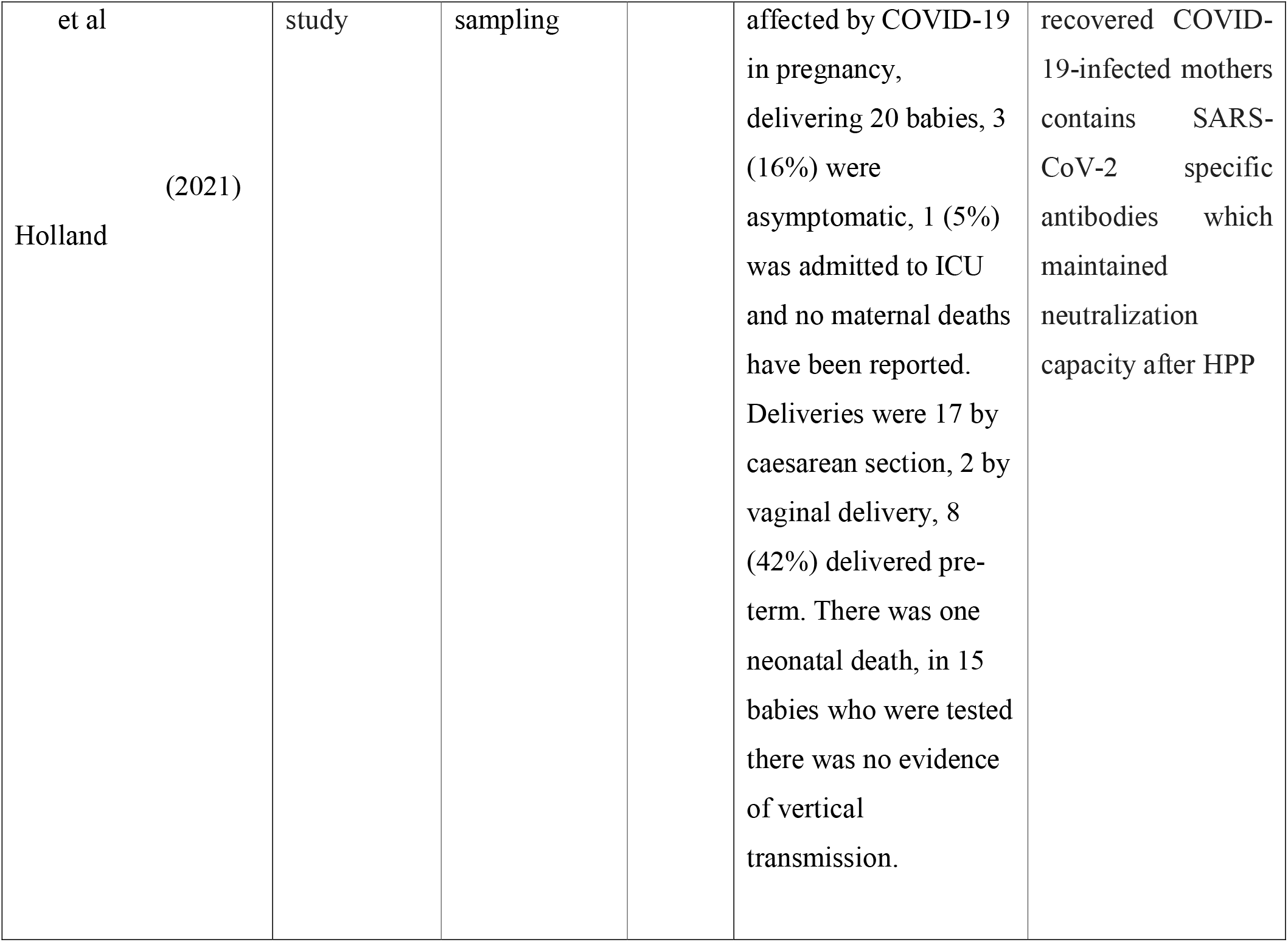
Descriptive Table.

**Table 4:**
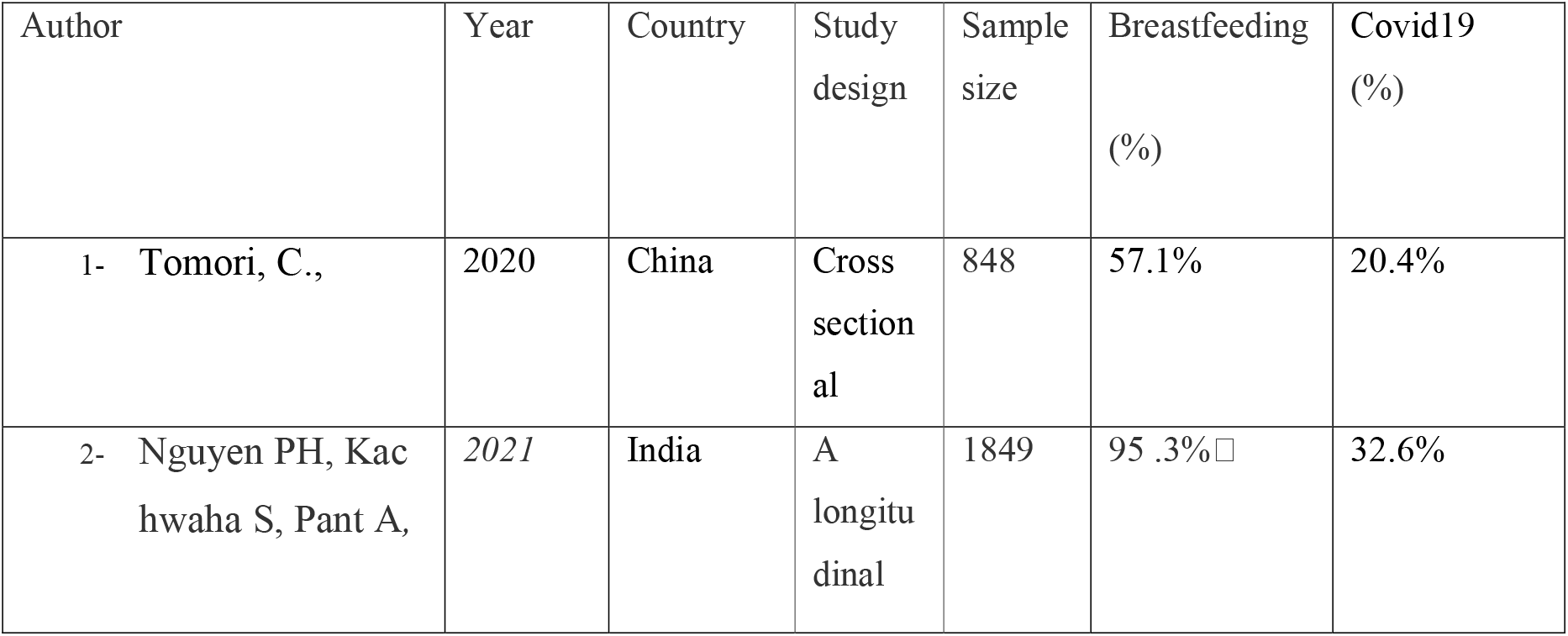
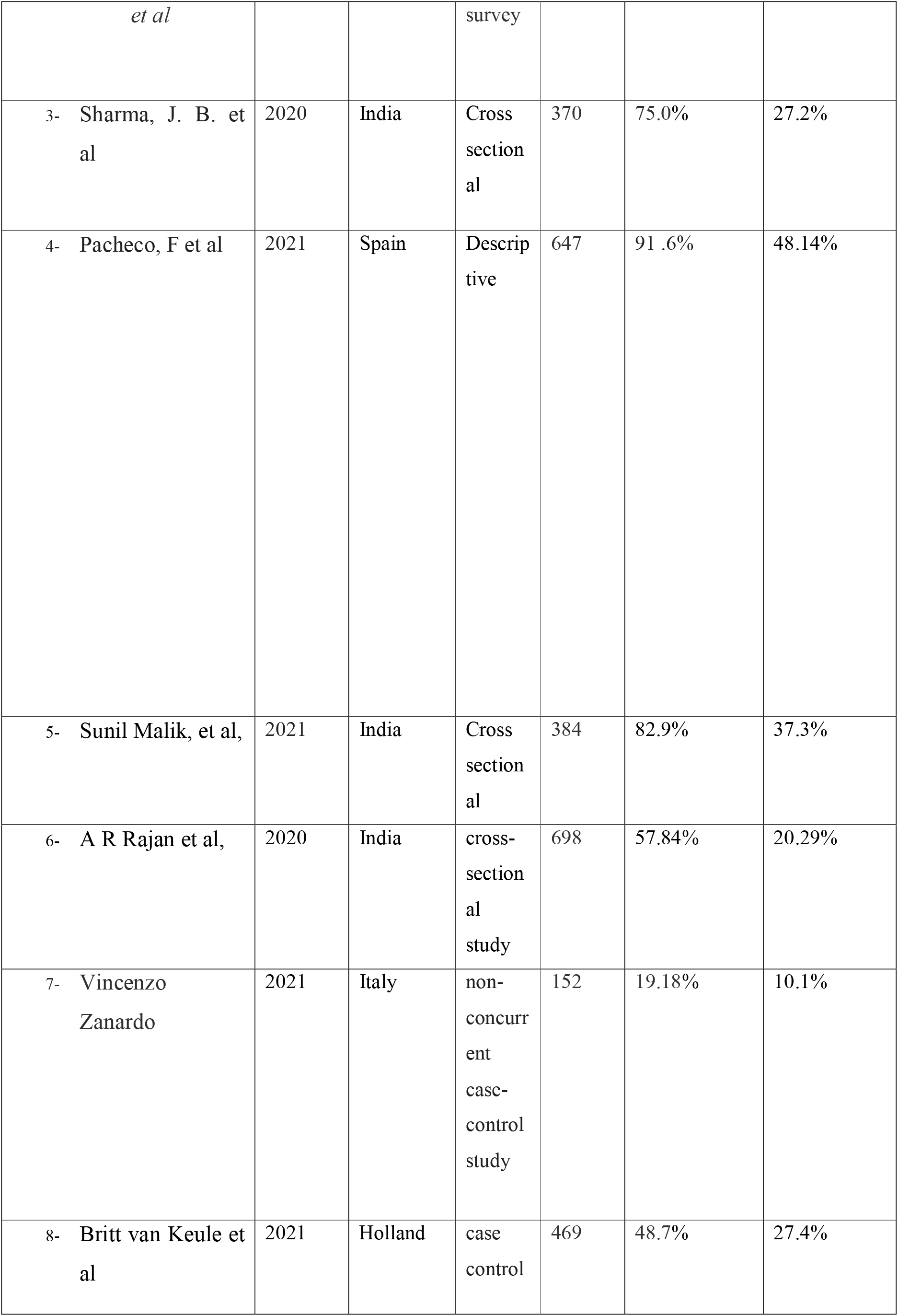
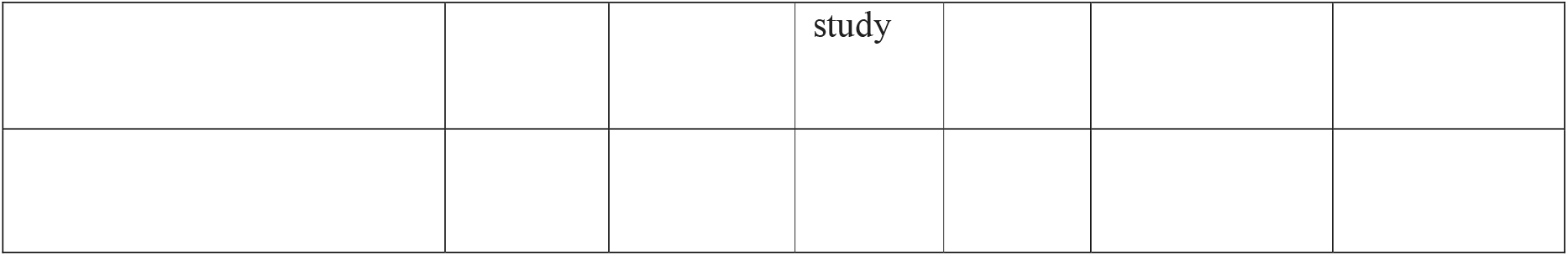
Systematic Review and Meta-Analysis.

### Quality assessment

Strobe checklist were used in this review to assess the quality of the articles reviewed. The strobe checklist is commonly used to critique and evaluate the quality of observational studies. The checklist consists of six scales which include: title, abstract, introduction, methods, results, and discussion these scales have subscales which results in total of 32 fields (subscales). In fact, these 32 fields represent different methodological aspects of a piece of research. example of subscales include title, problem statement, study objective, study type, statistical papulation, sampling method, sample size, the definition of variables and procedures. data collection method(s), statistical analysis techniques, and findings.

## Result

A total of 3160 records were identified in this systemic review with eight relevant studies involving 159 mothers (63 mothers with COVID-19 positive, 55 of their breast milk samples tested negative for the-Covid-19. Twenty-one breast milk samples from 8 women tested positive for Covid-19. Of 73 infants were born to mothers with COVID-19 at the time of delivery. Two infants tested positive for Covid-19. The average mother-child separation time was 36•7 ± 21•1 days among mothers confirmed with COVID-19. Out of 22 mothers, ((37.5%) chose to breastfeed their babies after confirm covid-19 positive.

## Discussion

This systemic review is on the breastfeeding practice among mothers during covid-19. This study has used all available secondary data of eight articles which is related these study and it examined thoroughly. All articles used in the study were different design. The aim was to focus on how covid-19 has impacted in breastfeeding practice during covid-19, the mental health status of mothers. The study shows that mothers have not changed their breastfeeding practices even confirmed positive cases and mother’s awareness about convid-19 were higher level. The study revealed that covid-19 has great impact on mothers mental health and frequency of daily breastfeeding, yet the degree of covid-19 transmission through breastmilk was low level. The most studies found that breastmilk is the best source of nutrition for new-born babies that can prevent infants to be effected by covid-19. It is because breastfeeding is an importance intervention to improve nutrition and health status infant child. It is however distribute of covid-19 in breastfeeding may leave negative impact on new-borns short and long-time health and also mothers. Therefore, an adequate breastfeeding practice among mothers during covid-19 is crucial for new-born health.

## Conclusion

Transmission of covid-19 from mothers to the new-born after birth, through breast milk and infectious respiratory was not reported. Much is still unknown about covid 19. There’s no recommendation to stop mother from their infants’ babies even if has been positively coronavirus confirmed. Current advice is to continue breastfeeding and to take all possible respiratory precautions and distancing to prevent spreading the virus to the infant. However, these recommendations may change with the rapidly evolving guidelines. Future studies on the protective functions and safety of breast milk during the coronavirus pandemic are needed.

## Data Availability

Data Availability

## Reference

1. Vijayalakshmi P, Susheelas T, Mythili D (2020). Knowledge, attitudes, and breast-feeding practices of postnatal mothers: A cross sectional survey. Int J Health Sci (Qassim).

2. Advisory for Human Resource Management of COVID-19. Advisory for Human Resource Management of COVID-19. [2020 Jun 16]; Available from: https://www.mohfw.gov.in/pdf/AdvisoryforHRmanagement.pdf

3. Chambers CD, Krogstad P, Bertrand K, Contreras D, Bode L, Tobin N, et al. Evaluation of SARS-CoV-2 in breastmilk from 18 infected women. medRxiv. 2020. https://doi.org/10.1101/2020.06.12.20127944.

4. Agampodi SB, Agampodi TC, Piyaseeli UKD (2020). Breastfeeding practices in a public health field practice area in Sri Lanka: a survival analysis. Int Breastfeeding J.

5. Sachdeva RC, Mondkar J, Shanbhag S, Sinha M, Khan A, Dasgupta R. (2019) A landscape analysis of human milk banks in India. Indian Pediatric

6. Sachdeva RC, Mondkar J, Shanbhag S, Sinha M, Khan A, Dasgupta R. (2019) A landscape analysis of human milk banks in India. Indian Pediatric

7. Ogbo FA, Agho KE, Page A 92015). Determinants of suboptimal breastfeeding practices in Nigeria: evidence from the 2008 demographic and health survey. BMC Public Health.

8. WHO Timeline - COVID-19. [Accessed cited 15 Jun 15, 2020]. Available from: https://www.who.int/newsroom/detail/27-04-2020-who-timeline.

9. Advisory for Human Resource Management of COVID-19. Advisory for Human Resource Management of COVID-19. [cited 2020 Jun 16]. Available from: https://www.mohfw.gov.in/pdf/AdvisoryforHRmanagement.pdf.

10. Stube, A. Should infants be separated from mothers with COVID-19? First, do no harm. Breastfeed. Med. 2020, 15, 351–352. [CrossRef] [PubMed].

11. Holcomb, J. Resisting guilt: Mothers’ breastfeeding intentions and formula use. Sociol. Focus 2017, 50, 361–374. [CrossRef].

12. WHO Guideline: protecting, promoting and supporting breastfeeding in facilities providing maternity and newborn services. Geneva: World Health Organization; 2017. Available from: https://www.who.int/nutrition/publications/guidelines/breastfeeding-facilities-maternity-newborn/en/.

13. ABM (Academy of Breastfeeding Medicine. ABM Statement on Coronavirus 2019 (COVID-19). 2020; 2020 Mar 10. Accessed: 2020 Mar 31. Available from: https://www.bfmed.org/abm-statement-coronavirus.

14. Schwartz DA. An Analysis of 38 Pregnant Women with COVID-19, Their Newborn Infants, and Maternal. Fatal Transmission of SARS-CoV-2: Maternal Coronavirus Infections and Pregnancy Outcomes. Arch Pathol Lab Med. 2020 Mar 17. [PMID: 32180426].

15. WHO. The optimal Duration of Exclusive Breastfeeding a Systematic Review; 2001. Accessed: 2020 Mar 30. Available from: https://www.who.int/nutrition/publications/optimal_duration_of_exc_bfeeding_review_eng.

